# Integrating ecological and anthropogenic risk identifies emerging Ebola spillover hotspots

**DOI:** 10.64898/2026.07.25.26358924

**Authors:** Monika Moir, Houriiyah Tegally, Desalew Meseret Moges, Graeme Dor, Jenicca Poongavanan, Cheryl Baxter, Richard J. Lessells, Moritz U.G. Kraemer, Ciara Judge, Bernardo Gutierrez, Dav. M. Ebengo, Eddy Kinganda-Lusamaki, Placide Mbala-Kingebeni, Jean-Jacques Muyembe-Tamfum, Jean B. Nachega, Tulio de Oliveira, Carla N. Mavian

## Abstract

The 2026 Bundibugyo virus outbreak emerged in a region with frequent conflict, food insecurity, rainforest and mining-related human mobility in Ituri province in the north-eastern region of the Democratic Republic of Congo (DRC). Existing ecological niche models have identified regions environmentally suitable for orthoebolavirus circulation but do not explicitly account for anthropogenic conditions that shape opportunities for interspecies contact, such as wildlife-to-human, and zoonotic spillover. Here, we update habitat suitability models for three putative orthoebolavirus reservoir bat species and for orthoebolavirus, and develop an integrated spatial spillover risk framework that combines ecological suitability with anthropogenic drivers, including human settlement, mining activity, bushmeat-related activities, forest loss, and conflict. We find that the updated model reveals previously under-predicted suitability in eastern DRC, and that the integration of anthropogenic factors with orthoebolavirus habitat suitability improves the prediction of historical zoonotic spillover locations. Boyce Index (measure of spatial predictive accuracy) increases from 0.78 to 0.97 when ecological suitability was combined with the built environment, while mining- and bushmeat-based scenarios showed the greatest enrichment of observed spillover events. We also find a temporal association of the relative contribution of habitat suitability and anthropogenic factors, with mining showing the largest and most consistent effect over the last decade, and conflict acting as a secondary amplifying factor whose apparent contribution has grown in recent periods. Together, these findings demonstrate that ecological suitability alone does not fully characterize landscapes vulnerable to Ebola zoonotic emergence and highlight the value of integrating environmental and anthropogenic information to strengthen One Health surveillance, epidemic preparedness, and targeted public health interventions in the DRC and neighboring countries.

## Main

Ebola virus disease (EVD) remains one of the most consequential zoonotic infectious diseases in Africa, causing recurrent outbreaks associated with high case-fatality rates, substantial disruption to health systems, and major socioeconomic consequences. The greatest public health burden has been caused by Ebola virus (formerly Zaire ebolavirus and referred to as *Orthoebolavirus zairense*), with reported case-fatality rates (CFR) ranging from approximately 40% to 90% across successive outbreaks, while Sudan virus (formerly Sudan ebolavirus) and Bundibugyo virus (Bundibugyo ebolavirus) have been associated with CFR of approximately 40-60% and 25-40%, respectively^1^. Although outbreaks are sustained through human-to-human transmission, they are typically initiated by zoonotic spillover from wildlife reservoirs ^2^. Identifying where spillover is most likely to occur is therefore fundamental to strengthening surveillance, preparedness, and early outbreak detection.

In early May 2026, a cluster of severe febrile illnesses affecting healthcare workers was detected in Bunia, Rwampara, and Mongbwalu Health Zones in the northeastern Democratic Republic of Congo (DRC) ^3–5^. Laboratory investigations subsequently identified Bundibugyo virus as the causative agent ^3^, confirming the third documented outbreak of Bundibugyo virus disease (BVD) worldwide, following outbreaks in Uganda (2007-2008) ^6^ and the Isiro region of northeastern DRC (2012) ^7^.

Substantial delays in laboratory confirmation and outbreak recognition are a notable epidemiological pattern that has emerged across Bundibugyo virus outbreaks. The 2007–2008 Uganda outbreak was characterised by approximately three months of unrecognised transmission before investigations were initiated ^6^. Similarly, retrospective molecular and epidemiological investigations of the 2012 Isiro outbreak in DRC revealed that Bundibugyo virus circulation had persisted for approximately 50 days before the previously recorded outbreak onset date, indicating substantial transmission prior to detection ^7^. Early investigations of the 2026 BDV outbreak indicate comparable delays, with evidence suggesting that transmission may have been ongoing for several weeks before laboratory confirmation ^8,9^. This diagnostic challenge stems partly from Bundibugyo’s lower case fatality rate ^10^, which makes outbreaks less epidemiologically conspicuous than those of the Zaire and Sudan Ebola species, and from the species-specificity of diagnostic assays, as commonly used rapid molecular diagnostics are designed to specifically detect *Orthoebolavirus zairense* virus ^11,12^.

Eastern DRC presents a uniquely complex setting for Ebola emergence, where ecological suitability for filovirus circulation intersects with social, environmental, and political instability. The region contains extensive forest ecosystems that harbour wildlife communities implicated in orthoebolavirus maintenance, while simultaneously undergoing rapid environmental transformation driven by population growth, extractive industries, and land-use change ^13^, creating conditions conducive to spillover from forest ecosystems to humans. Over the past three decades, recurrent armed conflict in North Kivu and Ituri provinces has created cycles of displacement, fractured healthcare infrastructure and compromised disease surveillance systems, creating conditions that have repeatedly hindered the control of Ebola outbreaks ^14^.

Since initial identification, the current BVD outbreak has expanded across multiple health zones in Ituri Province, neighbouring provinces, and spread into neighbouring Uganda, with one exported case in France. Response efforts face substantial challenges, including armed conflict, population displacement, healthcare-facility transmission, limited surveillance capacity, and the lack of approved vaccines and therapeutics, with a trial of the first treatments for BVD now underway ^15^. This vulnerability is not unique to the present crisis: the 2018-2020 Kivu Ebola outbreak persisted due to long-standing conflict, including violent attacks on treatment centres and healthcare teams, and recent resurgences have occurred in conflict-affected regions of Guinea and the DRC where outbreak control measures cannot be effectively implemented ^16,17^.

Beyond these direct effects on outbreak response, conflict ^18^ and displacement ^19^ in eastern DRC increasingly force communities to rely on artisanal mining, bushmeat hunting ^20^, and other forest-based livelihoods, intensifying human encroachment into fragmented wildlife habitats where contact with potential orthoebolaviruses reservoirs is more frequent ^21^. Artisanal mining settlements have been proposed to play a dual role in disease emergence: functioning both as ecological amplification zones characterised by environmental disturbance, high population mobility, overcrowding, and limited healthcare access; as well as strategic surveillance nodes where clusters of unexplained febrile illness may provide early warning of emerging infectious threats ^22^.

In recent years, eastern DRC has experienced the convergence of multiple public health and humanitarian crises, including the recent mpox Clade 1b outbreak ^23^ and cholera outbreaks ^24^, widespread food insecurity ^25^ and ongoing armed conflict that has displaced hundreds of thousands of people ^26^. These crises and extreme resource limitations have caused cascading vulnerabilities to infectious disease in this region. Displaced populations in the region have experienced disproportionately high burdens of cholera and other communicable diseases ^27^, reflecting highly constrained access to clean water, sanitation infrastructure, and healthcare services. The recent dismantling of displacement camps around Goma and subsequent population dispersal into informal settlements and rural areas has further disrupted access to healthcare, disease surveillance networks, and humanitarian assistance ^27^. Simultaneously, reductions in international aid and escalating insecurity have weakened already fragile health systems ^28,29^.

We hypothesise that these overlapping factors create a syndemic environment in which conflict, displacement, loss of livelihood, and human encroachment into fragmented forests interact synergistically to drive disease emergence and persistence. Under this framework, concurrent epidemics and humanitarian emergencies compete for limited healthcare resources and surveillance capacity while simultaneously increasing spillover risk by driving populations toward forest-based livelihoods and increasing reliance on wildlife resources. The resulting combination of heightened exposure to wildlife reservoirs and reduced access to healthcare may create ecological and social conditions that elevate the risk for orthoebolavirus spillover, human-to-human transmission and delayed outbreak recognition. Rather than viewing orthoebolavirus emergence solely as an ecological phenomenon, this syndemic hypothesis conceptualises emergence as the outcome of interacting environmental, anthropogenic, and epidemiological pressures operating across multiple spatial and temporal scales.

In this study, we investigate the sociodemographic and environmental correlates of historical Ebola spillover events and generate spatially explicit maps highlighting areas of elevated multifactorial spillover risk. Our approach provides a simple and interpretable framework for integrating ecological and human dimensions of spillover risk, with potential applications for targeting surveillance and preparedness activities in resource-limited settings. Identifying locations where these factors converge may improve the ability to anticipate and prevent future outbreaks before they escalate into larger public health emergencies.

### Updated bat distribution and Ebola ecological mapping models highlight previously underappreciated Ebola virus niche in Ituri Province

Identifying areas where ecological suitability for orthoebolavirus transmission coincides with human vulnerability remains a major challenge, and understanding the spatial distribution of reservoir animal species is central to achieving this. Fruit bats, particularly the hammer-headed bat (*Hypsignathus monstrosus*), Franquet’s epauletted fruit bat (*Epomops franqueti*), and little collared fruit bat (*Myonycteris torquata*), have been implicated as potential Ebola reservoirs ^30,31^. However, direct isolation of orthoebolaviruses from these species remains limited, and definitive reservoir hosts have not been identified ^32^. Previous ecological niche modelling studies have mapped the distribution of putative reservoir bat species ^33^, and predicted zoonotic transmission niche and spillover risk across Central and West Africa ^34–36^. These ecological suitability maps are widely recognised as risk maps for disease emergence. Notably, eastern DRC regions including Ituri province show only moderate predicted suitability for Ebola transmission risk ^35^. Recognising that the moderate predicted suitability in the eastern DRC does not reflect the region’s documented emergence history, we sought to update both the potential bat reservoirs and viral ecological suitability maps to better account for regional drivers of spillover.

By mapping the distribution of these fruit bat species, we identify areas of high ecological reservoir suitability that may serve as contact zones with displaced and forest-dependent populations. This is useful to distinguish areas with high ecological potential for spillover from areas where spillover has been realised, which occurs when ecological conditions are compounded by syndemic pressures. We mapped the relevant fruit bat species distributions (**Figure 1**) with an updated curation of occurrence points (**Supplementary Figure 1**) and an ensemble modelling approach with high predictive performance (*H*. *monstrosus* Area Under the Receiver Operating Characteristic Curve (AUC) > 0.89 and and True Skill Statistic (TSS) > 0.82; *E*. *franqueti* AUC > 0.88 and TSS > 0.71; *M. torquata* AUC > 0.87 and TSS > 0.73). Our results indicate the probability of occurrence for the three bat species south of the Sahara, from west to east Africa, including large tracts of Central Africa. The three species show consistently high suitability across most of West Africa, tracking the West African forest belt, extending from Sierra Leone to Cameroon. The habitat suitability of all three species also spans the Congo Basin rainforest region from Equatorial Guinea and Gabon, east through the northern areas of the DRC. The suitability map of *H*. *monstrosus* displays the highest suitability in the DRC. The inferred distribution of these bat species in our study is extended further east across Uganda and southeastern regions of Ethiopia compared to previously published maps ^33,34^.

**Figure 1:**
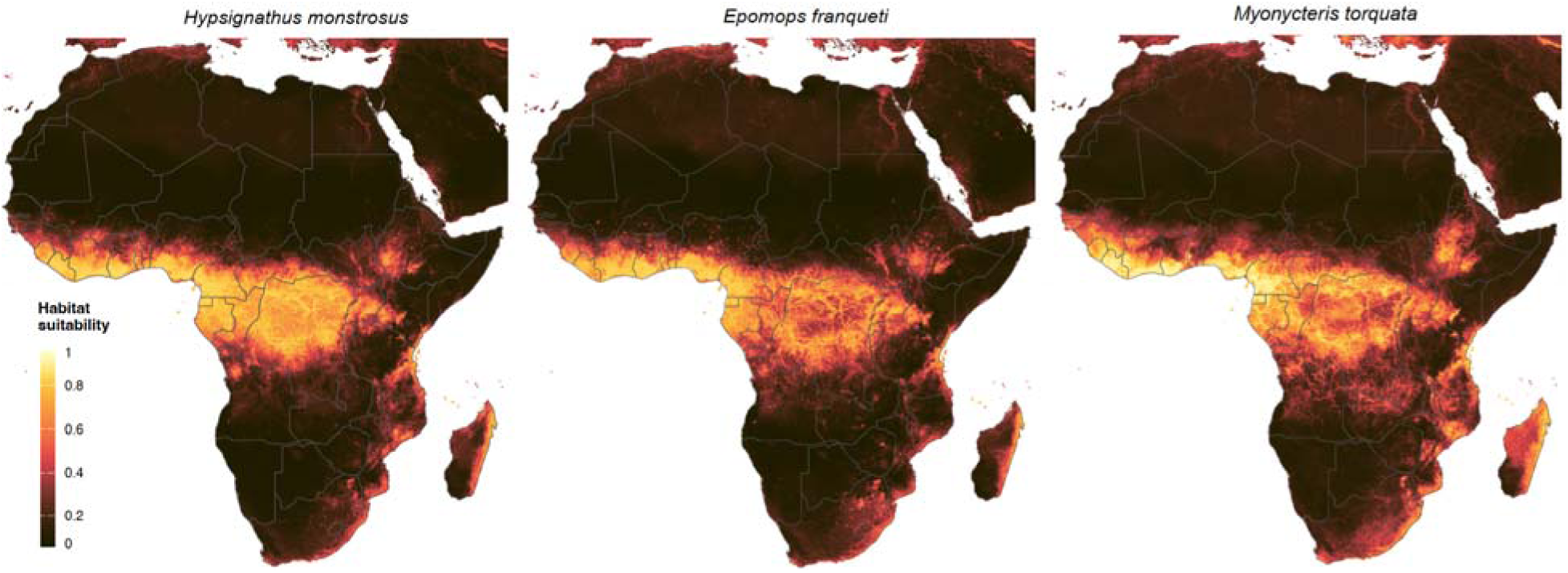
Predicted habitat suitability for three African fruit bat species that are putative orthoebolavirus reservoir hosts. Habitat suitability maps for *Hypsignathus monstrosus*, *Epomops franqueti*, and *Myonycteris torquata* across the African continent. The map shows country borders with grey polygons. The colour legend represents a scale of relative probability of occurrence in that location, from 0 (unsuitable in black) to 1 (highly suitable in yellow).

We followed a similar ensemble modelling procedure to update the predictions of environmental suitability for zoonotic transmission across Africa for all orthoebolavirus species. We expanded the viral occurrence dataset via a literature search following the methods and criteria of previous work ^34,35^. This dataset consisted of confirmed human index cases and animal infections from 1976 to 2026 (**Supplementary Figure 2**). We present the predicted zoonotic niche suitability using occurrence data through 2016, updated predictions incorporating records through 2026, and the difference map showing shifts in relative probability (**Figure 2**).

**Figure 2:**
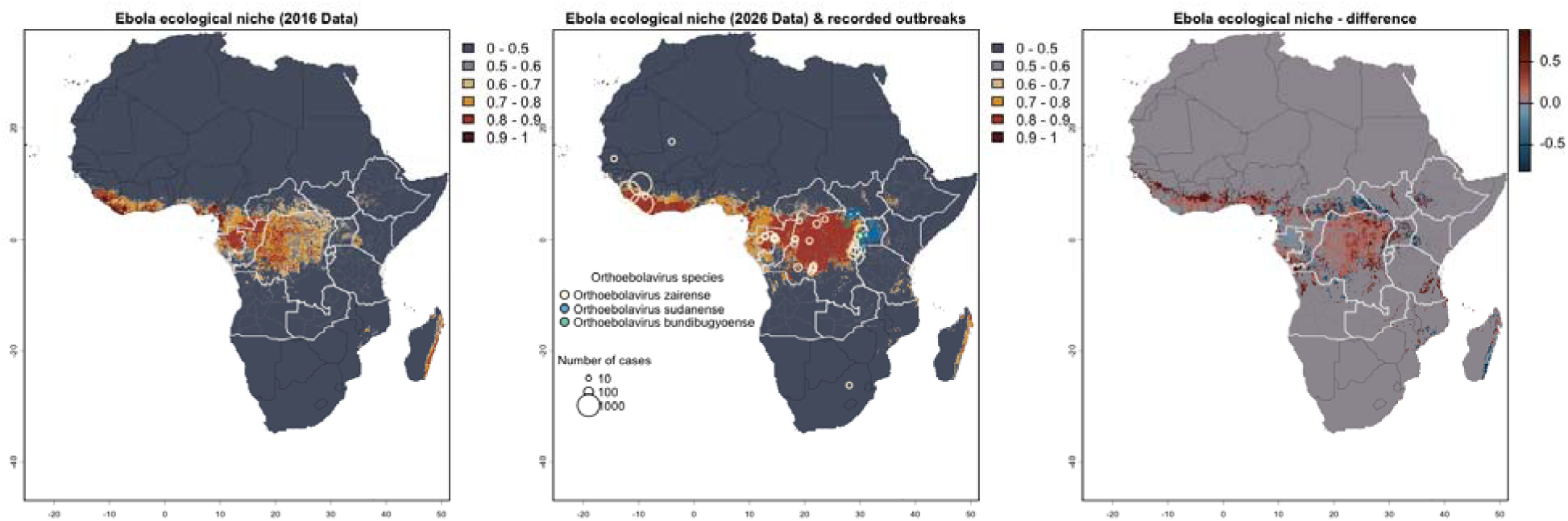
Updating the zoonotic niche of orthoebolavirus with a decade of emergence data. Predicted environmental suitability for orthoebolavirus spillover based on occurrence data from 1976 to 2016; and the updated prediction incorporating occurrence records up to 2026, including the current outbreak in the eastern DRC; and the difference map (2026 minus 2016) which highlights regions where predicted environmental suitability has increased (red) or decreased (blue), revealing how recent epidemiological data reshapes our understanding of Ebola spillover geography. The colour legend for the first two maps represents a scale of relative probability of occurrence from 0 (unsuitable in dark blue) to 1 (highly suitable in dark red). Countries visualized with white borders represent our study area for downstream analysis. Recorded outbreaks locations and sizes are shown on the middle panel, with circle colours representing respective orthoebolavirus species.

Our 2016-calibrated model broadly aligns with previously published maps^35^; however, we delineate a larger high-suitability region across West Africa and extend suitability into western DRC, whereas the cited analysis identifies central regions of DRC as highly suitable. Our updated 2026 model reveals substantially increased suitability in West Africa, particularly in Sierra Leone, Liberia, Côte d’Ivoire and Ghana. Additionally, there is expansion of predicted suitability through the Republic of Congo and northern DRC. Notably, the updated model assigned higher environmental suitability to eastern DRC, bringing predicted suitability into closer spatial agreement with recently documented orthoebolavirusspillover events in the region.

We used Shapley Additive Explanations (SHAP) values to quantify the mean importance and the SHAP-based marginal contribution of each environmental variable to individual predictions of the zoonotic niche suitability for both 2016- and 2026-calibrated models to understand differences between the two models (Supplementary Figure 3). Evapotranspiration (land-atmosphere water transfer) was the most influential predictor for the 2016 model (17.6%), and the Enhanced Vegetation Index (EVI; a measure of vegetation greenness and density) for the 2026 model (10.4%). We note that both evapotranspiration and EVI showed positive relationships with predicted suitability, whereby higher values of each variable were associated with positive SHAP contributions, indicating increased orthoebolavirus suitability. The differences in the importance of environmental variables between the two time periods may be due to changes in forest extent and vegetation condition, as they are proxies for canopy cover and vegetation productivity. We highlight reduced orthoebolavirus suitability along the northern and southern margins of the Congo Basin rainforest and increases in coastal Angola. This pattern corresponds with climate reconstructions showing decreased climate suitability for forest along these forest margins, while conditions in coastal Angola became increasingly favourable for forest vegetation ^37^.

### Syndemic vulnerability reveals predicted hotspots of orthoebolavirus spillover risk in eastern DRC

Although previous studies have identified areas environmentally suitable for orthoebolavirus circulation^33,35,36,38^, we have highlighted that these may not fully capture the anthropogenic factors that influence exposure and transmission. To address this gap, we adopted a pragmatic risk-mapping framework that combines orthoebolavirus ecological suitability with spatial indicators of human population presence, accessibility to reservoirs, and conflict-related vulnerability. By scaling ecological suitability according to these anthropogenic factors, we identify areas where environmental conditions conducive to virus circulation overlap with elevated potential for human exposure and disease spread.

We evaluated multiple spillover-risk scenarios by overlaying orthoebolavirus ecological suitability with anthropogenic drivers, including mining activity ^22^, conflict exposure ^14^, bushmeat hunting ^39^, forest loss ^40^, and built environment (**Figure 3**). We compared two versions of the orthoebolavirus ecological suitability map (model estimated with data curated up to 2016 ^35^, and model estimated with updated data curated up to 2026 in this paper) to assess whether updated risk mapping models improve predictive accuracy. We evaluated our spillover-risk maps (**Figure 4A**) for their ability to predict known locations of zoonotic orthoebolavirus occurrence (Supplementary Data/Table) for three time periods: <2010 to capture dynamics of orthoebolavirus emergence prior to the expansion of mining activities in DRC ^13,41,42^, 2010-2021 to capture a period characterised by increased mining and conflict but prior to intensified conflict events in the region, and ≥2022 to characterise a period of further intensification of conflict exposure in the region (**Supplementary Figure 4**). Additionally, since the early 2000s, the area affected by deforestation has quadrupled, while built-up areas have expanded by at least 100km^2^ every 5 years in the predicted Ebola niche in Central Africa (**Supplementary Figure 5**). For each scenario and time period (<2010, 2010-2021, and ≥2022), model performance was assessed using the Boyce Index and enrichment metrics. These metrics quantify the ability of risk surfaces to discriminate between observed spillover locations and the degree to which spillovers occur in areas of elevated predicted risk relative to the background landscape. While Boyce demonstrates how well a model can identify where spillovers are likely to occur, enrichment identifies how strongly spillovers are concentrated within the predicted high-risk areas.

**Figure 3.**
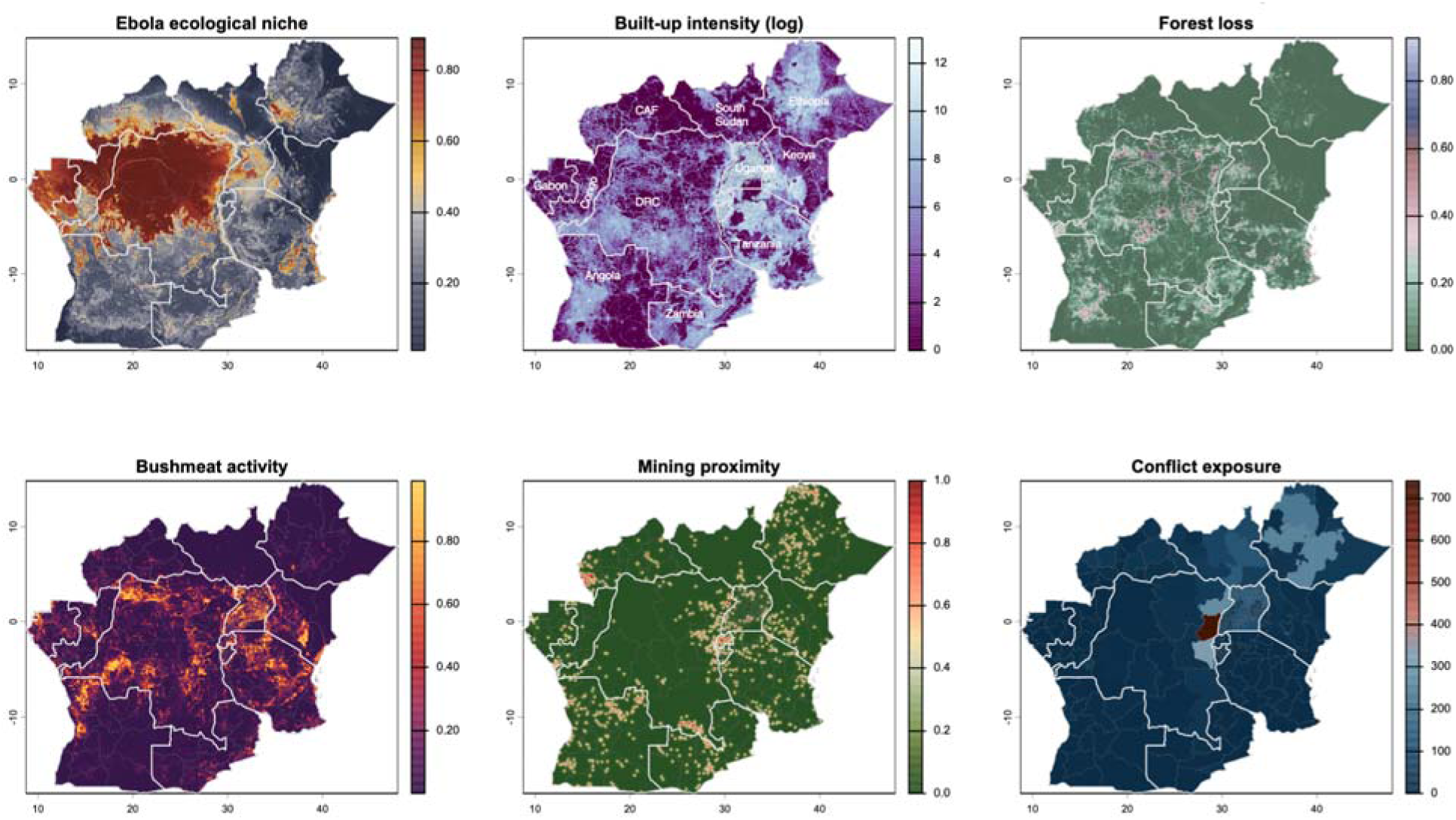
Spatial distribution of ecological and anthropogenic drivers of orthoebolavirus spillover risk across the DRC and neighbouring countries. Maps show six key variables integrated to predict spillover and transmission risk in eastern DRC. (Top left) Orthoebolavirus ecological niche: updated 2026 modeled habitat suitability for orthoebolaviruses circulation. (Top middle) Built-up intensity (log-transformed): Nighttime light-derived settlement intensity, quantifying human population concentration and urbanization patterns. (Top right) Forest loss: Proportion of forest cover loss between 2000-2025, indicating habitat fragmentation and land-use change that may alter human-wildlife contact patterns. (Bottom left) Bushmeat activity: Spatial density of bushmeat hunting and wildlife exploitation activity. (Bottom middle) Mining proximity: Distance-weighted proximity to active artisanal and large-scale mining sites; warmer colors indicate closer proximity to mining operations. (Bottom right) Conflict exposure: Conflict intensity (number of conflict events aggregated over 2000-2026) was derived from Armed Conflict Location and Event Data (ACLED), with darker colors indicating higher cumulative conflict exposure. Country names are annotated in the top-middle panel, except for Rwanda and Burundi.

**Figure 4.**
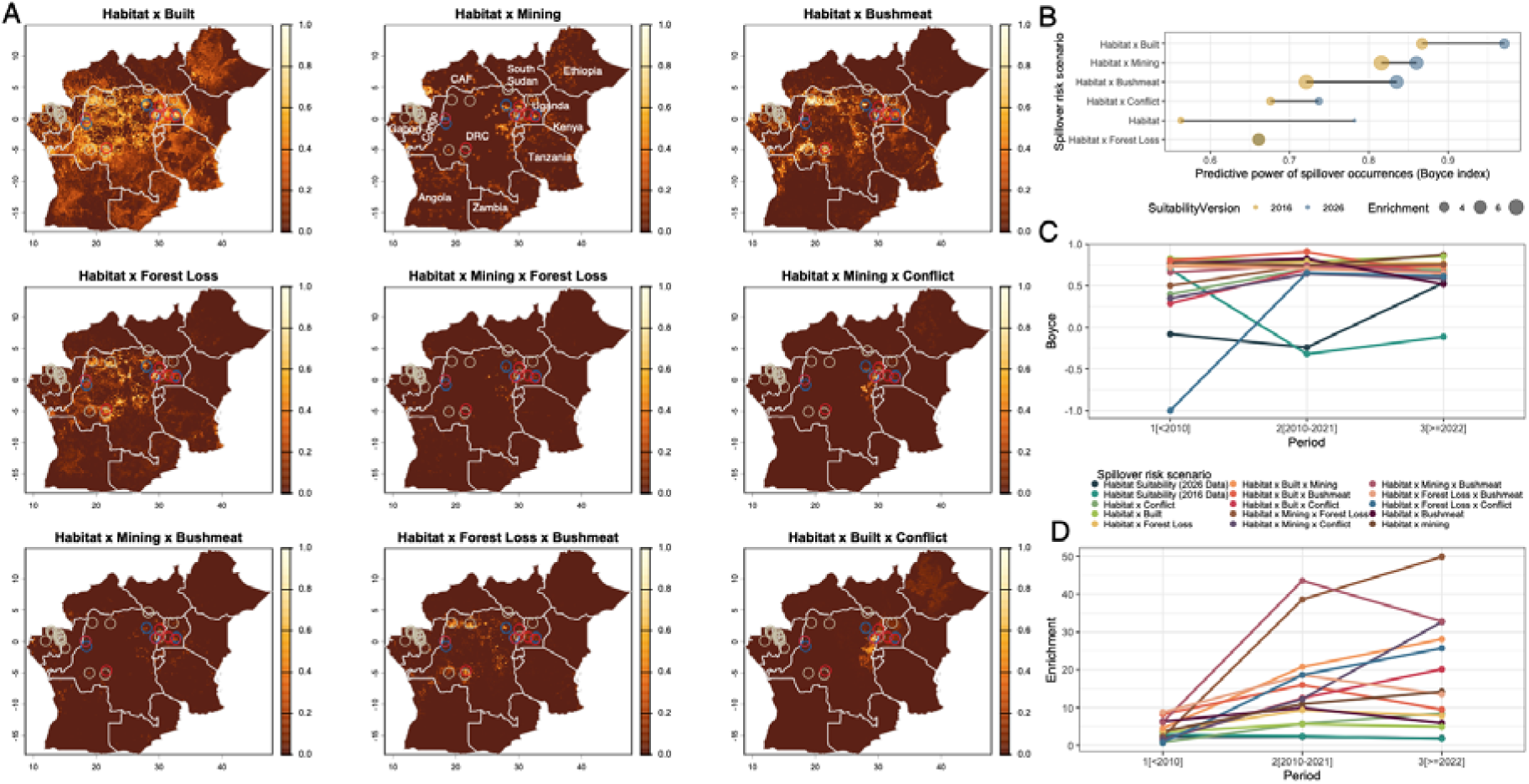
Integrating anthropogenic drivers with ecological suitability improves prediction of orthoebolaviruses spillover hotspots. **(A)** Spatial risk maps generated by combining the updated 2026 orthoebolavirus ecological suitability surface with anthropogenic drivers associated with spillover and transmission. Shown are representative composite risk scenarios integrating ecological suitability with built environment, mining proximity, bushmeat activity, forest loss, mining plus forest loss, mining plus conflict, mining plus bushmeat, forest loss plus bushmeat, and built environment plus conflict. Risk values are scaled from 0 to 1, with brighter colors indicating greater predicted spillover risk. Circles indicate historical zoonotic orthoebolavirus spillover locations used for model evaluation, coloured by date (White: <2010; blue: 2010-2021; red: ≥2022). Country names are annotated in the top-middle panel, except for Rwanda and Burundi. **(B)** Predictive performance of each spillover-risk scenario measured using the Boyce Index. Points represent models generated using the 2016 (yellow) and updated 2026 (blue) orthoebolavirusecological suitability surfaces, with point size proportional to enrichment of observed spillover events within predicted high-risk areas. **(C)** Temporal comparison of model performance (Boyce Index) across three periods (<2010, 2010-2021, and ≥2022), illustrating changing contributions of anthropogenic drivers to spillover prediction over time. **(D)** Corresponding enrichment values for each spillover-risk scenario across the three time periods.

**Figure 5.**
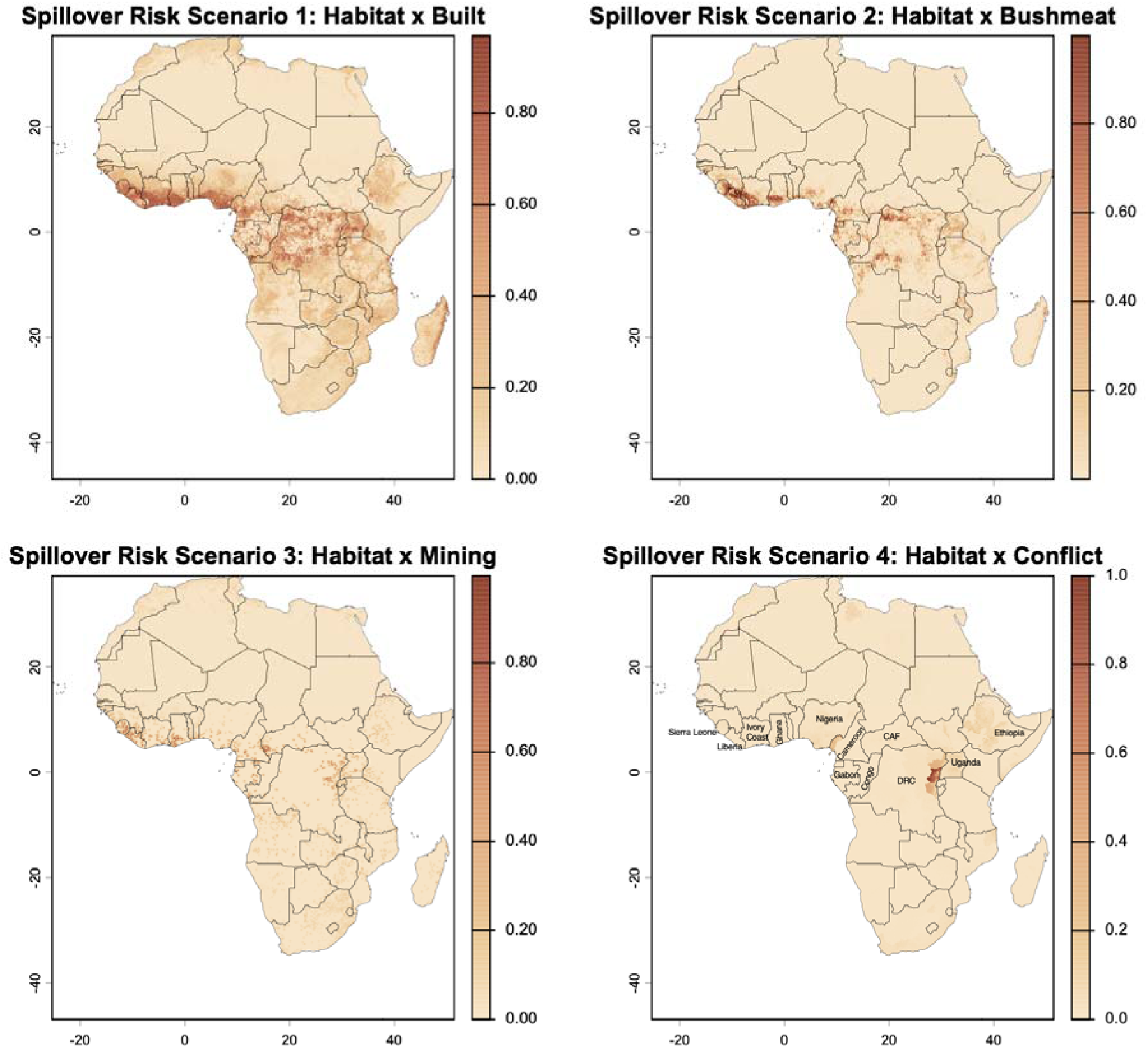
Continental Ebola spillover risk map integrating ecological habitat suitability with the four principal anthropogenic drivers identified in upstream analysis: built environment, bushmeat activity, mining proximity, and conflict exposure. The composite risk surface highlights areas across Africa where suitable ecological conditions overlap with anthropogenic pressures, identifying potential spillover hotspots.

We find that orthoebolavirus habitat suitability alone demonstrated moderate predictive performance, with the updated 2026 suitability model outperforming the 2016 model (Boyce = 0.78 vs. 0.56) (**Figure 4B**). This indicates an improved ability of the updated model to characterize spillover landscapes. Incorporating anthropogenic drivers consistently enhanced model performance across all scenarios. Built environment yielded the greatest improvement (Boyce = 0.97), suggesting that spillover events are concentrated at the interface between suitable habitats and human-modified landscapes. Mining and bushmeat exposure substantially increased enrichment (6.8-8.3-fold and 6.8-7.8-fold, respectively), highlighting the importance of extractive activities and wildlife exploitation in shaping spillover hotspots. In contrast, conflict events and forest loss produced more modest gains, suggesting that they may act as secondary amplifying factors rather than primary drivers of spillover. Overall, these findings indicate that while ecological suitability defines the underlying spillover landscape, anthropogenic pressures actively concentrate spillover risk within specific high-risk environments. Importantly, while the model performance for the majority of tested scenarios improves with the updated habitat suitability model, intersecting orthoebolavirus ecological suitability improves predictive accuracy even with the 2016 model. This highlights the continued relevance of previously published risk mapping work for epidemic preparedness when considered within a multi-risk framework.

Temporal analysis of the spillover-risk maps reveals how the importance of anthropogenic drivers shifted across the three distinct time periods. Prior to 2010, models incorporating habitat suitability with built environment, bushmeat exposure, and forest loss showed the strongest performance (Boyce = 0.71-0.83), while habitat suitability alone performed poorly (**Figure 4C**). During 2010-2021, combinations of habitat suitability with bushmeat, mining, and built environment achieved the highest discriminatory ability, with the habitat-built-bushmeat model performing best overall (Boyce = 0.91), and mining-related models exhibiting markedly elevated enrichment values (**Figure 4C,D**). After 2022, models integrating habitat suitability with built environment, mining, forest loss, and conflict continued to perform strongly, with habitat-built, habitat-mining-forest loss, and habitat-built-conflict scenarios achieving the highest Boyce indices and enrichment values (**Figure 4C,D**). This recent period reflects intensifying regional conflict and environmental disruption from mining. Collectively, these findings suggest that while ecological suitability provides the underlying landscape for spillover potential, the spatial concentration of recent spillover events is increasingly shaped by interactions between suitable habitats and temporally shifting anthropogenic landscape modification, including mining activity, human settlement, forest disturbance, wildlife exploitation, and exposure to conflict.

### Continental risk maps based on four main anthropogenic drivers identify broader hotspot locations

Building on the spillover risk scenarios that best improved spillover occurrence predictions, we generated continental-scale risk maps by integrating orthoebolavirus habitat suitability with the four principal anthropogenic drivers identified in our analyses: mining activity, bushmeat exposure, built environment, and conflict intensity. These maps highlight regions where favourable ecological conditions coincide with different human pressures at the continental scale, enabling the identification of potential spillover hotspots beyond historically affected areas. Such risk maps can facilitate preparedness in countries at high risk of anthropogenic-driven spillovers by supporting targeted surveillance, wildlife monitoring, and public sensitization around risky practices. We find key regional differences in risk related to the four individual factors. For instance, while built-up environment helps to narrow estimated risk from environmental suitability to areas with human settlement across most of the orthoebolavirus ecological niche, bushmeat-related activity concentrates spillover risk in northern and the western part of DRC, in almost all of Liberia, and in parts of Sierra Leone, Ivory Coast, Ghana, Nigeria, Cameroon, and Uganda. According to these risk maps, mining could drive zoonotic spillovers in Sierra Leone, Liberia, Ghana, Central African Republic, parts of Cameroon, Equatorial Guinea, and Gabon, and around DRC. Finally, eastern DRC, and to a lesser extent Ethiopia and Cameroon are highlighted for being at risk of conflict-related modulation of spillover risk.

## Discussion

Our study demonstrates that integrating ecological suitability with anthropogenic drivers substantially improves prediction of Ebola spillover hotspots compared with ecological suitability alone. Using updated environmental suitability models and spatial data on mining, bushmeat activity, built environment, forest loss, and conflict, we identified areas where ecological conditions conducive to orthoebolaviruses circulation intersect with human activities that increase opportunities for spillover. Notably, areas identified as having the highest composite risk showed substantial overlap with locations affected by the current outbreak. While our work does not establish causality, it supports the biological and epidemiological relevance of the framework and suggests that integrating ecological and sociodemographic factors may improve the identification of areas vulnerable to Ebola emergence and spread.

Our findings further suggest that conflict is an important component of Ebola risk landscapes in eastern DRC. Prolonged insecurity can increase human exposure to wildlife reservoirs through population movement, livelihood disruption, and environmental disturbance, while simultaneously weakening surveillance systems, limiting healthcare access, and delaying outbreak detection and response. These interacting pressures can facilitate both zoonotic spillover and onward transmission. Incorporating dynamic information on population displacement and healthcare accessibility into future spatial risk models could further improve prediction of disease spread and support more targeted preparedness and response strategies.

An important implication of our findings is that Ebola spillover risk cannot be adequately represented by a single continent-wide model. While ecological suitability defines where reservoir hosts and favorable environmental conditions occur, our analyses demonstrate that the anthropogenic processes triggering spillover vary substantially between regions. This was similarly seen in a multicriteria risk-mapping study at a landscape scale in Guinea, Congo, and Gabon, which found the relative contributions of environmental, bushmeat-related and reservoir species risk factors to Ebola spillover differed between eco-regions, even when forest cover and human population density were comparable ^36^. However, this study ^36^ showed forest cover and forest loss as the most sensitive risk factors for spillover risk at a landscape scale, while we found forest loss to have modest gains in predictive performance at a broader scale. This difference may reflect scale-dependence in the relative contributions of various drivers, highlighting the need for eco-regional risk assessments.

Our analysis showed that in Central Africa, human settlement, bushmeat activity and mining emerged as dominant predictors, likely compounding the effects of environmental disturbance, increased human encroachment into wildlife habitats, population mobility and reduced access to healthcare. In contrast, other regions in Africa may be driven primarily by agricultural expansion, deforestation or different forms of environmental disturbance. These findings suggest that spillover prediction should move from static ecological niche models towards region-specific frameworks that integrate the local social, environmental, and ecological context. Such an approach would provide more accurate and operationally relevant risk maps, allowing surveillance resources to be directed towards the human-environment interfaces where spillover is most likely to occur. This may also inform surveillance strategies that can explicitly account for the heterogeneous anthropogenic pressures that shape pathogen emergence in different landscapes.

More broadly, this work addresses an important gap in understanding the interaction between environmental change, conflict, forced displacement, and Ebola emergence. The approach has potential operational relevance for preparedness and surveillance activities in the DRC and neighbouring countries, particularly where resources for monitoring and response are limited. By identifying locations where ecological suitability coincides with human vulnerability, risk assessments can be used to support prioritization of surveillance, community engagement, vaccination strategies, and healthcare resource allocation. These findings have immediate implications for One Health surveillance.

A notable strength of the framework is that it achieves substantial improvements in predictive performance using a pragmatic and readily reproducible modelling strategy. Rather than applying complex weighted interaction models, all predictors were combined using a direct multiplicative overlay. This approach minimizes subjective assumptions regarding the relative importance of individual drivers, facilitates interpretation, and is readily reproducible across pathogens and geographic regions. Given the limited number of confirmed zoonotic spillover events available for model calibration, more complex approaches may increase the risk of overfitting and reduce model generalizability. Although the framework is intentionally simple and should not be interpreted as a mechanistic model of spillover, it demonstrates the value of integrating ecological and human dimensions of risk within a common spatial framework. Similar approaches could be adapted to other zoonotic and epidemic-prone pathogens, including Marburg virus disease, mpox, cholera, and other emerging infectious diseases whose transmission dynamics are influenced by environmental change and population vulnerability. As such, these findings contribute to ongoing efforts to strengthen epidemic intelligence, One Health surveillance, and anticipatory approaches to outbreak preparedness in regions facing multiple interacting ecological and societal pressures.

This work must be interpreted in light of several limitations. First, while the locations of spillover events used in our analysis have been carefully curated with literature context, they represent only the best approximation of actual zoonotic exposure, as human infections can be detected after delayed case detection and movement of affected individuals. Second, the relatively small number of spillovers (n=48) may affect statistical power and evaluation metrics, particularly in the further-stratified temporal analysis. Additionally, our framework does not account for the risk of Ebola resurgence from viral reservoirs in human survivors. This transmission pathway is mechanistically distinct from zoonotic spillover and whose contribution to landscape-level risk cannot be estimated within our current approach; although outbreaks from new zoonotic spillovers have been the majority ^43^. Third, due to data accessibility constraints, the anthropogenic drivers were treated as static or aggregated throughout the time periods, which is likely an oversimplification, potentially underestimating the temporal effects of human interaction with the environment. Consequently, temporal comparisons primarily reflect changes in the spatial distribution of spillover events relative to current landscapes, and future work incorporating time-varying anthropogenic datasets may further refine these relationships. In terms of determining the impact of deforestation on zoonotic spillovers, the spatial resolution of our analysis may underestimate small-scale forest loss, particularly in the DRC where smallholder agricultural plots are perhaps the primary drivers of deforestation; re-estimating forest loss intensities from higher spatial resolution satellite imagery, which is outside the scope of the current study, would address this limitation. Finally, additional factors not considered here likely affect both the likelihood of zoonotic spillover and onward spread. These include, for instance, information about displaced populations and healthcare accessibility of recipient communities, which can be integrated in a future framework investigating the trajectory of realised spillovers. For future work, it would also be important to consider that it is highly likely that different orthoebolavirus species could have different niches, although this is so far unresolvable given the gaps in knowledge related to reservoir hosts.

In conclusion, our findings demonstrate Ebola spillover as an emergent property of interacting ecological and anthropogenic processes, rather than a function of environmental suitability alone. By coupling updated reservoir and viral niche models with spatially explicit indicators of mining, conflict, bushmeat hunting, and land-use change, we show that the landscapes most vulnerable to spillover are those where ecological conditions and human activity converge, and that the relative contribution of these anthropogenic pressures changes as regional crises evolve. The spatial overlap between our risk surfaces and the ongoing Bundibugyo virus outbreak in the DRC lends practical support to this framework. As the DRC and ecologically and politically comparable regions continue to face pressures from conflict, displacement, and environmental change, we suggest that integrative region-specific risk frameworks offer a pragmatic tool for One Health surveillance that can be rapidly updated as new data become available.

## Methods

### Fruit bat ecological suitability modelling

Ecological niche models were developed for three fruit bat species (*Hypsignathus monstrosus*, *Epomops franqueti*, and *Myonycteris torquata*) using the biomod2 ensemble modelling framework in R ^44^. Occurrence records were downloaded from the Global Biodiversity Information Facility (GBIF) ^45–47^ and updated with records from the most recent African Chiroptera Report ^48^. Records were cleaned and spatially thinned to match the raster of environmental variables at ∼10 km^2^. Following previously published methodology ^33^, models were calibrated using environmental predictors comprising bioclimatic variables from WorldClim 2.1 ^49^ with a spatial resolution of 5 arc-minutes, the coefficient of variation of the enhanced vegetation index (EVI) from EarthEnv ^50^ compiled from 16-day composites collected between 2001 and 2005, and the 2021 ESA WorldCover land cover fractional layers ^51^. All environmental predictors were resampled to 5 arc-minutes. Environmental variables were screened for multicollinearity using pairwise correlation coefficients (r > 0.7) and a variance inflation factor (VIF) threshold of 10, resulting in a final set of 13 predictors retained for modelling (BIO5, 6, 13, 14 and 15, EVI, and land cover classes: trees, grassland, shrubs, cropland, built, water, and wetland). For each bat species, an ensemble of algorithms (combinations of Generalized Linear Models (GLM), Gradient Boosting Machines (GBM), Random Forests (RF), Generalized Additive Model (GAM), and Maximum Entropy (MAXNET)) was fitted with the weighted mean prediction, with only individual models achieving a True Skill Statistic (TSS) ≥ 0.7 retained for ensemble construction. Three pseudo-absence replicates were sampled using a disk strategy (150–1000km from presences) at a 1:1 presence to pseudo-absence ratio, evaluated using Area Under the Receiver Operating Characteristic Curve (AUCroc) and True Skill Statistic (TSS) metrics with 3-repeat 5-fold cross-validation, and final predictions were projected across Africa. Maps were plotted in R.

### Suitability mapping of the zoonotic niche of Ebola virus disease

We employed ensemble species distribution modelling to map environmental suitability of Ebola zoonotic spillover events across Africa. We utilised the Ebola occurrence dataset from a previously published study^35^ and updated the records of confirmed animal infections via a literature search of PubMed, from 2015 to 2026, following the methodology outlined in published record^34^. We sourced updated human index case locations from previous work ^52^. Occurrence data comprised 93 confirmed Ebola detections spanning 1976 to 2026. A total of 89 georeferenced occurrence records were used alongside 178 pseudo-absence points generated within the accessible area (M region). We define the M region as all ecoregions sharing biomes with documented Ebola occurrence records, following the BAM framework ^53^. Pseudo-absence points were constrained to locations greater than 150 km from any presence record to ensure they represented ecologically similar but epidemiologically distinct regions.

Following previous work ^34^, we compiled the following environmental covariates (at 5 km resolution): Enhanced Vegetation Index (EVI) mean and range from MODIS monthly data ^54^, mean and range of land surface temperature from ERA5 daily aggregates 2010–2025 ^55^, SRTM (CGIAR-CSI processed) elevation ^56^, mean evapotranspiration ^57^. Environmental variables were screened for multicollinearity using a combined correlation coefficient and VIF approach described above. The final set of environmental predictors retained for modelling were mean EVI, mean LST daytime, LST range, elevation, and mean evapotranspiration. We implemented four algorithms within the R biomod2 ^44^ framework: GLM, GBM, RF, and MAXNET. Models were evaluated using 5-fold cross-validation repeated three times, with performance assessed using AUC and TSS. An ensemble projection was constructed from all models with TSS > 0.8. Model predictions were projected across the African continent to identify geographic areas of heightened zoonotic niche suitability.

### Geospatial data for spillover risk modelling

#### Spillover occurrence data

Our spillover occurrence dataset consisted of updated human index case locations from previous work ^52^, consisting of 48 occurrence records from 1976 to 2026, which we geocoded to the nearest available known location in published reports.

#### Built-up environment data

We obtained data on built-up environment globally for the years 2000-2025 (5-year intervals), from Global Human Settlement Layer Dataset ^58^ at 30 arcsec resolution. We then cropped each layer to the relevant study area. For modelling simplicity, we selected the 2025 layer for multiplicative risk estimation described below to represent the most recent conditions. We log transformed the built-up raster layer using the *log1p* function in R. Since we are assessing potential interaction of ecological suitability with anthropogenic activity, spillover risk resulting from areas with small populations are worth considering; this guided our choice to log-transform the built-up environment raster layer.

#### Mining data

Mining-site locations were compiled primarily from OpenStreetMap (OSM), using features tagged as mining-related infrastructure (e.g industrial=mine, landuse=quarry, man_made=mineshaft, man_made=adit, historic=mine, and abandoned or disused quarry tags). Where available, additional non-OSM artisanal/small-scale mining records were incorporated from open mining-site datasets, including the Rwanda ICGLR mining-site database ^59^.

All mining-site records were converted to point locations using the mapped point coordinate or the centroid of the mapped OSM geometry. Sites were then rasterized to a regular 0.05-degree (30km2) grid covering Africa. For each grid cell, we calculated the distance to the nearest rasterized mining-site cell using a Euclidean distance transform. Distances were converted from grid-cell units to kilometers using latitude-adjusted degree lengths. The resulting raster represents distance, in kilometers, to the nearest known mining-related site in the compiled dataset.

This was then transformed to a mining proximity metric, modelled as an exponentially decreasing function of distance to the nearest mining site: *e^−D/k^*, where *D* is the distance to the nearest mining site and *k=10* represents a 10-km scale of spatial decay.

#### Conflict events and exposure data

Conflict data were obtained from the Armed Conflict Location & Event Data (ACLED) project ^60^, a widely used geospatial database that provides near real-time information on violence, demonstrations, and associated population exposure. Two complementary conflict indicators were extracted: (i) conflict events, representing the occurrence of individual incidents such as battles, violence against civilians, riots, and protests; and (ii) population exposure, representing the estimated number of people residing within a 5 km radius of each recorded conflict event, derived using WorldPop population estimates. We aggregated ACLED records to first-level administrative units (Admin-1) for three study periods (<2010, 2010–2021, and >2022) and calculated mean annual conflict events and population exposure. These estimates were then converted into continuous raster layers at approximately 5 km resolution and used as spatial predictors in subsequent analyses.

#### Bushmeat activity data

Bushmeat activity data was obtained from previously published modelled estimates ^61^.

#### Forest loss data

Forest loss data (tiles) were obtained for Africa from the Global Forest Change dataset (https://glad.earthengine.app/view/global-forest-change), which provides annual global tree cover loss estimates at 30 m spatial resolution based on Landsat satellite observations ( ^62^; updated annually; data obtained for 2000-2025). Raster tiles were processed individually by converting forest loss pixels into a binary disturbance layer (1 = forest loss, 0 = no forest loss) and aggregated to a coarser spatial resolution to facilitate integration with other environmental predictors. Processed tiles were mosaicked into a continuous Africa-wide raster, with overlapping cells averaged to generate a harmonized forest loss surface.

### Spillover risk modelling framework

A spatial spillover-risk framework was developed by integrating ecological suitability with anthropogenic drivers hypothesised to influence pathogen spillover. For this analysis, we considered a study area comprising DRC and neighbouring countries (Republic of Congo, Uganda, Rwanda, South Sudan, Burundi, Tanzania, Angola, Central African Republic, Zambia, Kenya, Gabon and Ethiopia). Habitat suitability surfaces were combined with raster layers representing mining activity, bushmeat activity, conflict intensity, built-up environment, and forest loss to generate spillover-risk scenarios. Prior to integration, all raster layers were projected to a common coordinate reference system (based on the Ebola ecological suitability layer), aligned to the same spatial resolution and extent, and resampled to a common analysis grid to ensure pixel-wise correspondence. We consider two versions of the Ebola ecological suitability map (model estimated with data curated up to 2016 ^35^, and model estimated with updated data curated up to 2026 in this paper) in order to determine the importance of updated risk mapping models for epidemic preparedness.

Composite spillover-risk maps were generated by combining habitat suitability with one or more anthropogenic layers using pixel-wise raster multiplication with prior normalisation of each individual raster layer. This approach assumes that spillover risk is greatest where favourable ecological conditions and anthropogenic exposure coincide, while areas with low values for any component contribute proportionally less to overall risk. Multiple scenarios representing different ecological hypotheses were evaluated, including habitat suitability alone, habitat combined with individual anthropogenic drivers, and habitat combined with multiple interacting drivers. The resulting composite risk surfaces were rescaled to a common 0-1 range using min-max normalization and subsequently evaluated against observed spillover locations using presence-only performance metrics.

To evaluate the ability of spillover-risk scenarios to identify observed spillover locations, each risk surface was assessed using the Boyce Index and an enrichment metric. Separate evaluations were performed for different time periods (<2010, 2010-2021, and ≥2022), corresponding to testing against zoonotic Ebola occurrence locations during those time periods to investigate temporal changes in the drivers of spillover risk. These time periods were chosen to capture shifting dynamics in the hypothesized anthropogenic variables: <2010 to capture dynamics of Ebola emergence prior to the amplification of mining activities in DRC ^41,42^, 2010-2021 to capture a period characterised by increased mining and conflict but prior to M23-related conflict events in the region, and ≥2022 to characterise a period of further intensification of conflict exposure in the region (Supplementary Figure 4).

Observed spillover locations were represented as georeferenced point locations and overlaid on each raster risk surface. For each scenario, predicted risk values were extracted at all spillover locations and compared with the distribution of risk values across the entire study area.

The Boyce Index was calculated using the *ecospat* implementation of the continuous Boyce Index in R ^63,64^, a presence-only evaluation metric widely used for ecological niche and habitat suitability models. The Boyce Index quantifies the extent to which observed spillover events occur in areas predicted to have higher risk than expected under a random distribution. Values range from −1 to 1, where positive values indicate that spillover events are preferentially associated with higher predicted risk, values close to zero indicate performance no better than random, and negative values indicate predictions that are inversely related to observed spillover locations. Boyce index values greater than approximately 0.6 were interpreted as indicating good discriminatory performance.

To quantify the concentration of spillover events within predicted hotspots, an enrichment ratio was calculated as:

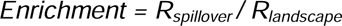

where *R_spillover_* is the mean predicted risk value extracted at observed spillover locations, and *R_landscape_* is the mean predicted risk across all raster cells in the study area.

An enrichment value greater than one indicates that spillover events occur disproportionately within areas of elevated predicted risk, while larger values reflect increasingly concentrated spillover hotspots.

## Data Availability

All model estimates produced in the present study are available upon reasonable request to the authors, and in preparation for public repository. All other data used in the study were already publicly available and all details are described within the manuscript.

## Supplementary Information

**Supplementary Figure 1:**
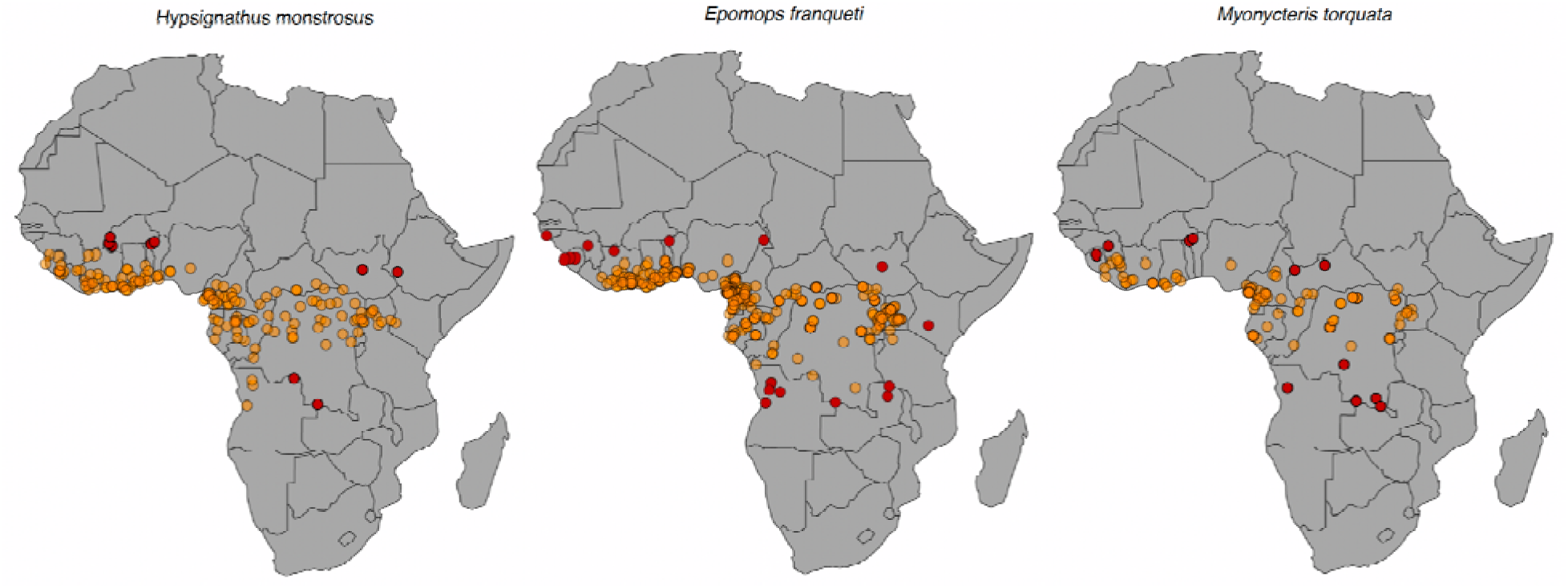
Spatial distribution of occurrence records for three putative Ebola reservoir bat species. Orange points denote occurrence records available from the Global Biodiversity Information Facility (GBIF) and the 2025 African Chiroptera Report. Red points highlight newly compiled records extending beyond the geographic scope of ^35^.

**Supplementary Figure 2:**
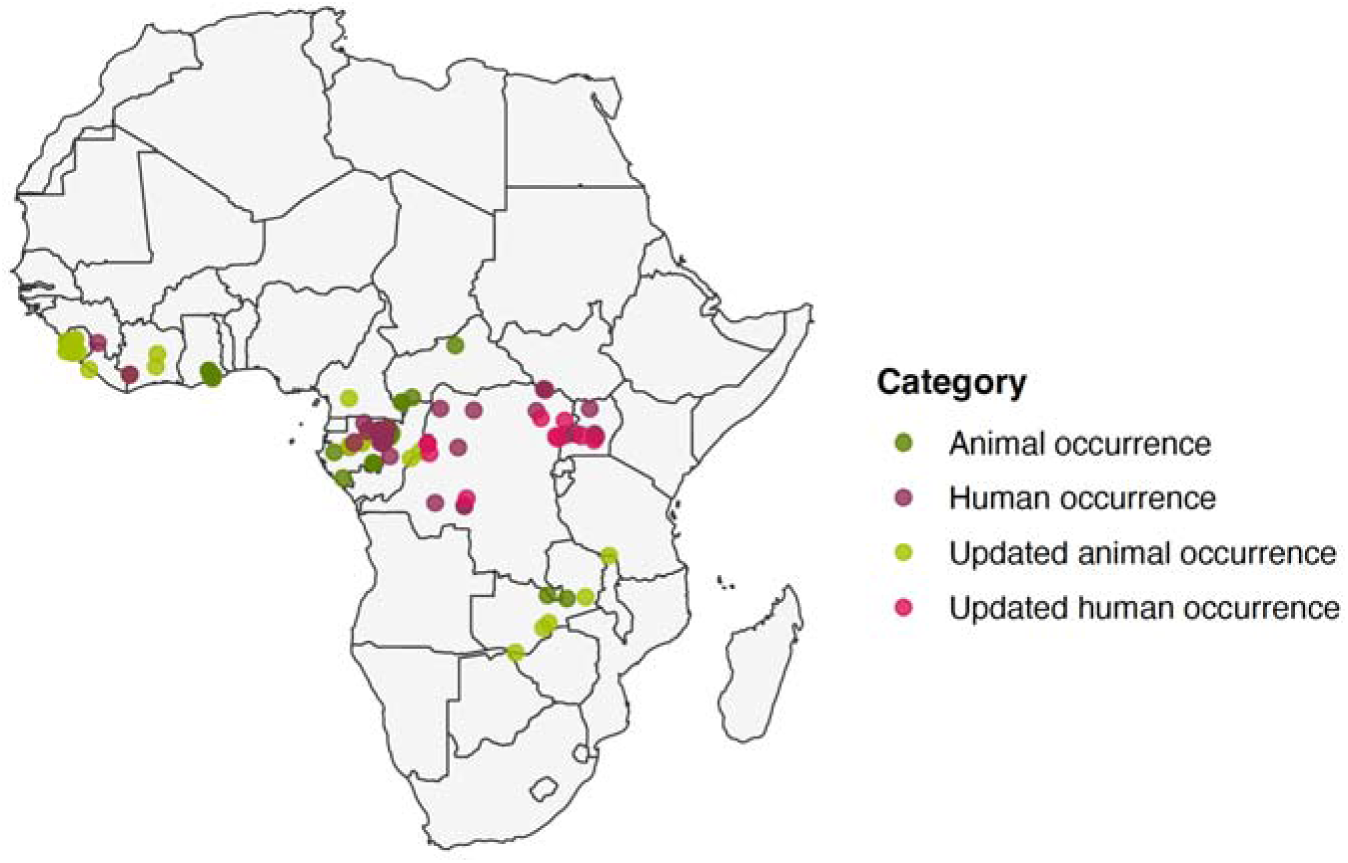
Updated African orthoebolavirus occurrence dataset (1976-2026). Confirmed human and animal orthoebolavirus occurrences compiled from a systematic literature review. Dark pink points show previously documented human index cases^35^; bright pink points represent newly compiled human occurrences. Similarly, dark green points show previously documented animal infections; bright green points highlight newly compiled animal occurrence records. The expanded dataset reflects updated surveillance records and newly identified spillover events through 2026.

**Supplementary Figure 3.**
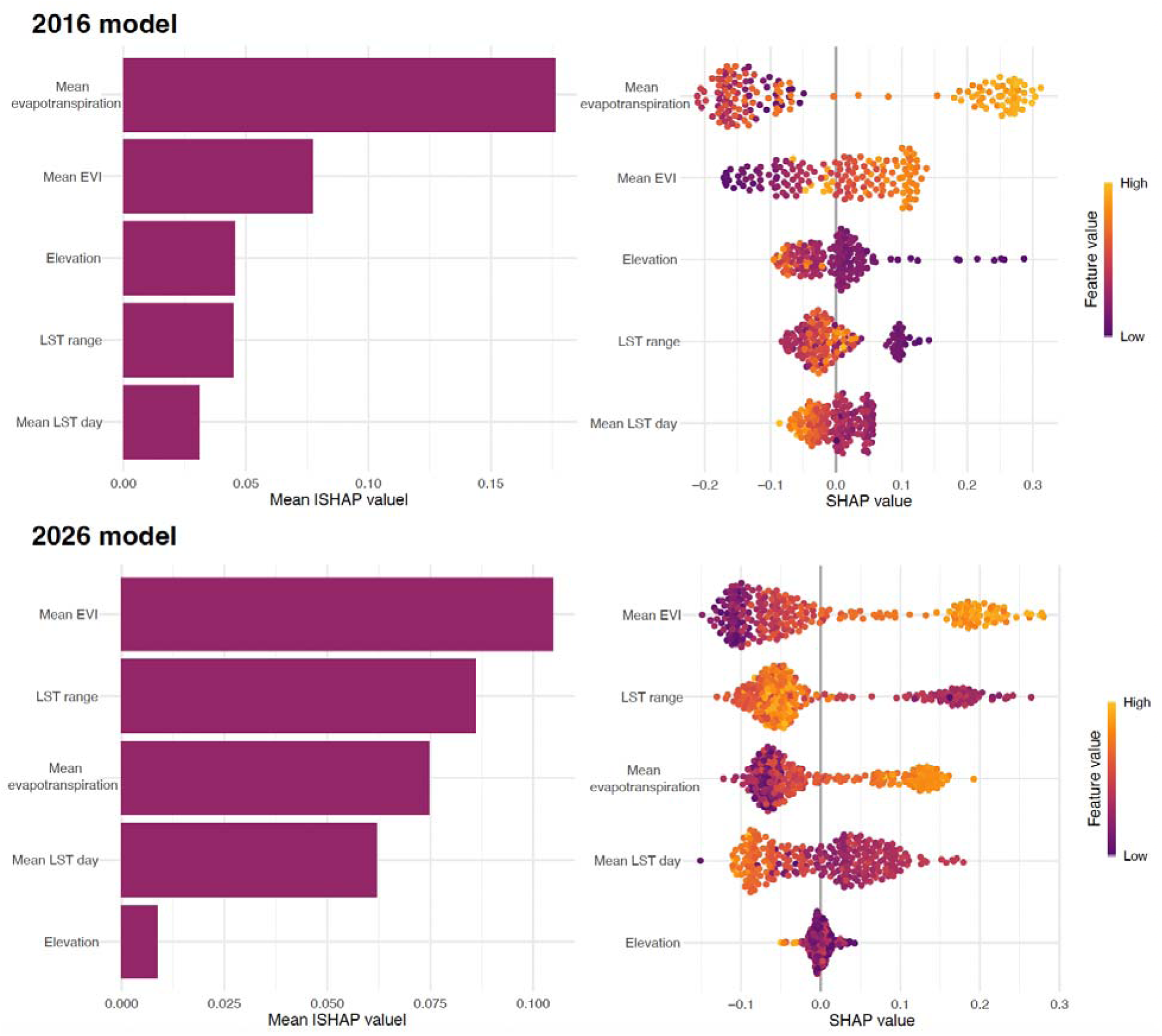
Environmental variable importance in orthoebolavirus zoonotic niche models. The top left figure shows the 2016-model, and the bottom left the 2026-model, variable importance calculated as the average of absolute SHAP values in the training dataset, indicating the average variable contribution to the overall prediction of the model. The beeswarm plot on the top right shows the distribution of SHAP values per variable for the 2016-model and the bottom right plot for the 2026-model. Beeswarm plots indicate whether environmental variable contributions are positive or negative, with low feature value coloured purple and high feature value in yellow.

**Supplementary Figure 4.**
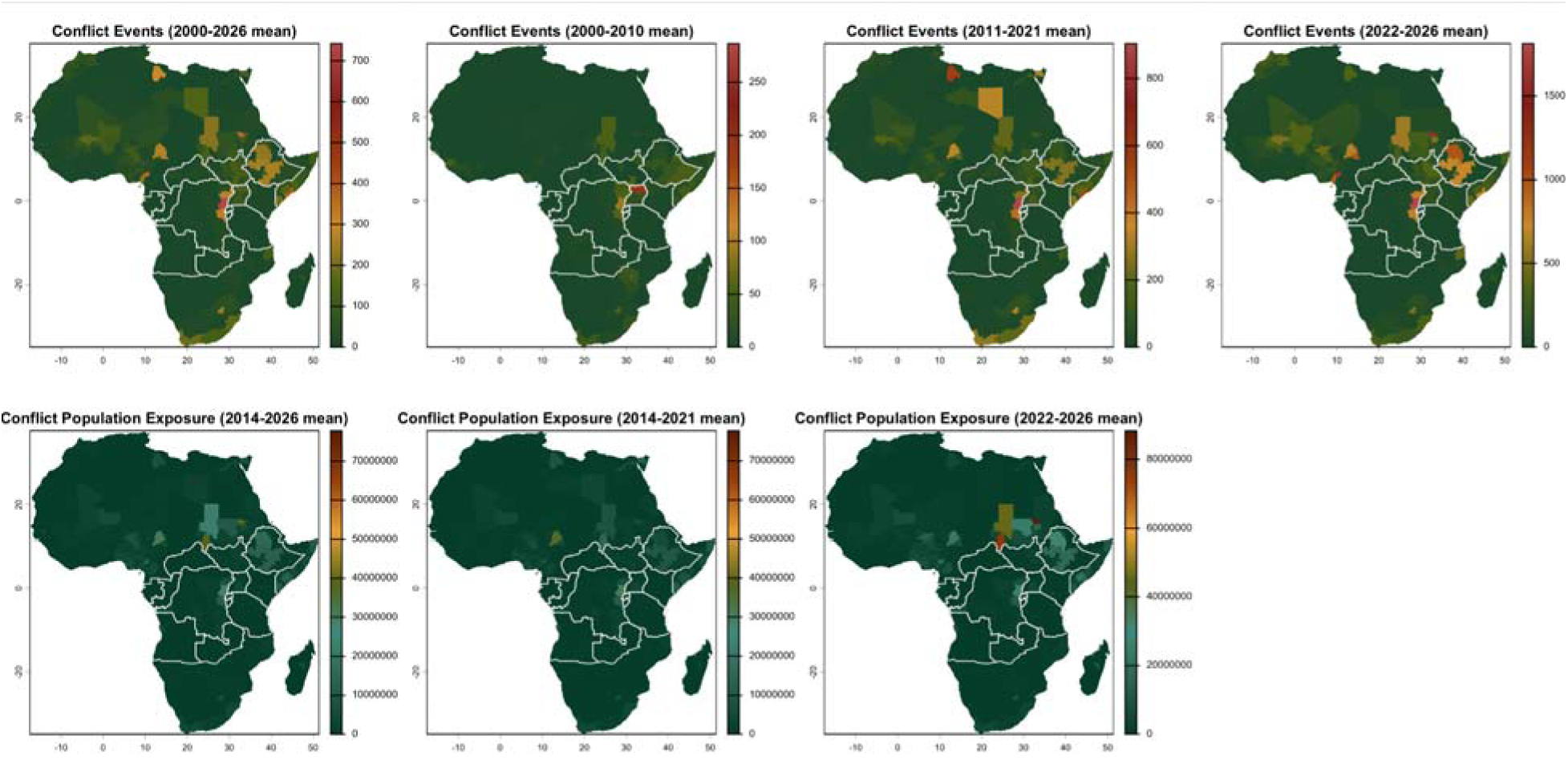
(Bottom row) Number of people exposed to conflicts; (Top row) Number of conflict events; both from 2000-2026.

**Supplementary Figure 5.**
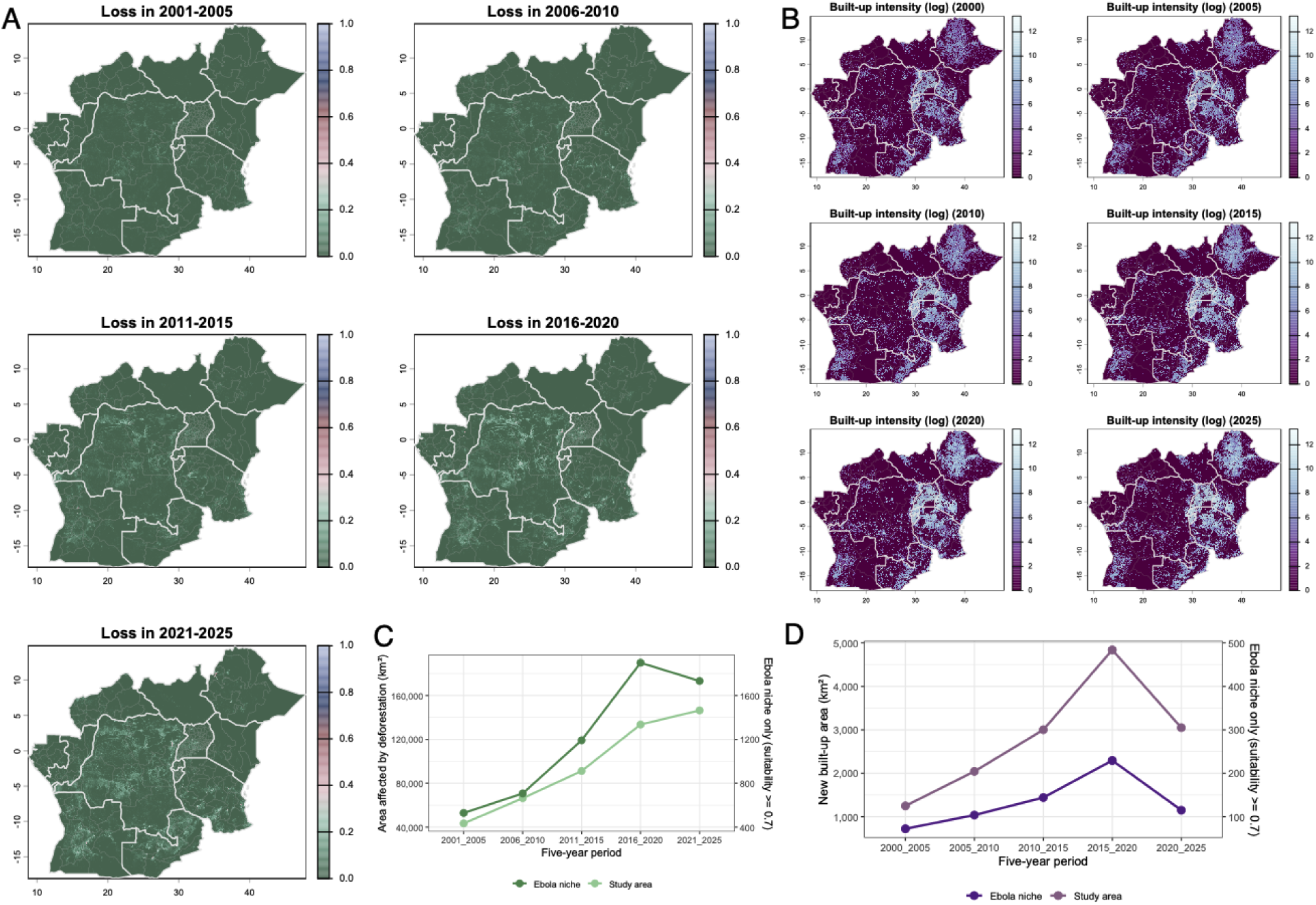
Temporal dynamics of forest loss and built-up expansion within the study area from 2000 to 2025. Maps show the spatial distribution of forest loss (A) and built-up expansion (B) across five-year internals within the study area. The line graphs show the corresponding temporal trends in the total area affected by forest loss (C) and built-up expansion (D) in km^2^ across the study region, with an additional focus on areas with Ebola ecological niche suitability >= 0.7.

## Data and code availability

All analyses were conducted on the freely accessible datasets, detailed within the methodology. Ecological suitability and spillover risk maps computed from this study are available online along with all codes to reproduce the analyses (https://github.com/CERI-KRISP/XXXX.git).

## Acknowledgements and funding statement

The authors acknowledge that this work is an initiative of the CLIMADE consortium (https://climade.health/). Research activities at CERI are supported in part by grants from the Rockefeller Foundation (HTH 017), the Abbott Pandemic DefenseCoalition (APDC), the SAMRC South African mRNA Vaccine Consortium (SAMVAC), Global Health EDCTP3 Joint Undertaking and its members as well as Bill & Melinda Gates Foundation (101103171), the UK’s Medical Research Foundation (MRF-RG-ICCH-2022-100069), the Wellcome Trust for the Global.health project (228186/Z/23/Z) and H.T.’s Career Development Award (335374/Z/25/Z), the Foreign, Commonwealth & Development Office (FCDO) (301489-405) and the Novo Nordisk Foundation (NNF24OC0094346). This research was also supported by the South African Medical Research Council with funds received from the South African National Department of Health and the UKRI Medical Research Council, with funds received from the UK Government’s International Science Partnerships Fund (grant 96881). G.D. acknowledges support from the SAMRC through its Division of Research Capacity Development under the SAMRC Clinician Researcher Development Programme, funded by the South African National Department of Health. M.U.G.K. acknowledges funding from The Rockefeller Foundation (PC-2022-POP-005), Health AI Programme from Google.org, the Oxford Martin School Programmes in Pandemic Genomics & Digital Pandemic Preparedness, European Union’s Horizon Europe project E4Warning (#101086640), Wellcome Trust grants 303666/Z/23/Z, 226052/Z/22/Z, the United Kingdom Research and Innovation (#APP8583), UK International Development (301542-403), the Bill & Melinda Gates Foundation grants (INV-063472, INV-090281), and Novo Nordisk Foundation (NNF24OC0094346). The contents of this publication are the sole responsibility of the authors and do not necessarily reflect the views of the European Commission or the other funders. D.M.E.,& P.M.K wish to thank the European Union – Horizon Europe / EDCTP3 Joint Undertaking for supporting this work through the EBOLA PREP TBOX project (Grant Agreement No. 101145709). D.M.E. P.M.K. & M.U.G.K wish to thank the Gates Foundation (INV-090281).

## Author contributions

H.T. and C.N.M. conceptualized, designed and supervised the study. M.M., H.T., D.M.M., J.P., G.D., and C.N.M., analysed data and did the primary data visualisations. M.M., H.T., and C.N.M. wrote the original draft. R.J.L, R.G. C.B., T.d.O., J.N., P.M., J.J.M., C.J. and M.U.G.K interpreted data. H.T., C.B., T.d.O acquired funding for this project. All authors reviewed and edited the final draft. All authors had final responsibility for the decision to submit for publication.

## References

1. Centers for Disease Control and Prevention (CDC). Ebola Disease Outbreak in the Democratic Republic of the Congo and Uganda. Health Alert Network https://www.cdc.gov/han/php/notices/han00530.html.

2. Feldmann, H., Sprecher, A. & Geisbert, T. W. Ebola. N. Engl. J. Med. 382, 1832–1842 (2020).

3. Centers for Disease Control and Prevention (CDC). Ebola Outbreak: Current Situation. Ebola https://www.cdc.gov/ebola/situation-summary/index.html#cdc_situation_summary_more_about-what-to-know-about-the-outbreak.

4. Mbulayi, O. et al. Real-time epidemic intelligence in a public health emergency: the 2026 Bundibugyo virus outbreak. Lancet Infect. Dis. (2026) doi:10.1016/S1473-3099(26)00330-0.

5. Institut National de Santé Publique et al. DRC Ebola Bundibugyo 2026. https://inrb-umie.github.io/BDBV2026-Epidemic_Dashboard/.

6. Wamala, J. F. et al. Ebola hemorrhagic fever associated with novel virus strain, Uganda, 2007-2008. Emerging Infect. Dis. 16, 1087–1092 (2010).

7. Hulseberg, C. E. et al. Molecular analysis of the 2012 Bundibugyo virus disease outbreak. Cell Rep. Med. 2, 100351 (2021).

8. Caelers, D. Month-long detection gap complicates Ebola containment in DRC. *Nat*. Africa (2026) doi:10.1038/d44148-026-00130-y.

9. Virological. Genomic epidemiology of the ongoing 2026 Bundibugyo Virus Disease outbreak in the Democratic Republic of the Congo - Bundibugyo ebolavirus - Virological. https://virological.org/t/genomic-epidemiology-of-the-ongoing-2026-bundibugyo-virus-disease-outbreak-in-the-democratic-republic-of-the-congo/1045.

10. Mwamba, D. et al. Bundibugyo virus disease outbreak in Ituri, Democratic Republic of the Congo. Lancet 407, 2367–2369 (2026).

11. Nsawotebba, A. et al. Clinical profile and genomic characterization of the 2026 Bundibugyo virus index case in Uganda. Nat. Med. (2026) doi:10.1038/s41591-026-04510-7.

12. MacNeil, A. et al. Filovirus outbreak detection and surveillance: lessons from Bundibugyo. J. Infect. Dis. 204 Suppl 3, S761–7 (2011).

13. Ladewig, M., Angelsen, A., Masolele, R. N. & Chervier, C. Deforestation triggered by artisanal mining in eastern Democratic Republic of the Congo. Nat. Sustain. 7, 1452–1460 (2024).

14. Kraemer, M. U. G. et al. Dynamics of conflict during the Ebola outbreak in the Democratic Republic of the Congo 2018-2019. BMC Med. 18, 113 (2020).

15. University of Oxford, Medical Sciences Division. Patient enrolment begins in PARTNERS trial to identify the first effective treatments for Bundibugyo virus disease. https://www.medsci.ox.ac.uk/news/patient-enrolment-begins-in-partners-trial-to-identify-the-first-effective-treatments-for-bundibugyo-virus-disease.

16. Charnley, G. E. C., Green, N., Kelman, I., Malembaka, E. B. & Gaythorpe, K. A. M. Evaluating the risk of conflict on recent Ebola outbreaks in Guinea and the Democratic Republic of the Congo. BMC Public Health 24, 860 (2024).

17. Shears, P. & Garavan, C. The 2018/19 Ebola epidemic the Democratic Republic of the Congo (DRC): epidemiology, outbreak control, and conflict. Infection Prevention in Practice 2, 100038 (2020).

18. Marou, V. et al. The impact of conflict on infectious disease: a systematic literature review. Confl. Health 18, 27 (2024).

19. Braam, D. H., Jephcott, F. L. & Wood, J. L. N. Identifying the research gap of zoonotic disease in displacement: a systematic review. Glob. Health Res. Policy 6, 25 (2021).

20. Spira, C., Kirkby, A., Kujirakwinja, D. & Plumptre, A. J. The socio-economics of artisanal mining and bushmeat hunting around protected areas: Kahuzi–Biega National Park and Itombwe Nature Reserve, eastern Democratic Republic of Congo. Oryx 53, 136–144 (2019).

21. Kavulikirwa, O. K. Intersecting realities: Exploring the nexus between armed conflicts in eastern Democratic Republic of the Congo and Global Health. One Health 19, 100849 (2024).

22. Sow, B. et al. Mining corridors as emergence interfaces: an evidence-informed one health operational framework for prevention of infectious disease outbreaks in Kédougou, Senegal. Front. Public Health 14, 1836395 (2026).

23. World Health Organization. Multi-country outbreak of mpox, External situation report #66 - 31 May 2026. https://www.who.int/publications/m/item/multi-country-outbreak-of-mpox--external-situation-report--66---31-may-2026 (2026).

24. Alam, M. T. et al. Emergence and Evolutionary Response of Vibrio cholerae to Novel Bacteriophage, Democratic Republic of the Congo1. Emerging Infect. Dis. 28, 2482–2490 (2022).

25. World Food Programme. Eastern DRC Ebola outbreak threatens to deepen hunger as WFP ramps up emergency support. https://www.wfp.org/stories/eastern-drc-ebola-outbreak-threatens-deepen-hunger-wfp-ramps-emergency-support (2026).

26. Bepouka, B., Kafua, M., Matangi, B., Longokolo, M. & Situakibanza, H. Simultaneous outbreaks of Ebola, cholera, mpox, and measles in DR Congo in 2025. Lancet 406, 2214–2215 (2025).

27. Maisha, F. M. et al. Cholera risk in Goma, DR Congo, after forced clearing of relief camps. Lancet 405, 1658–1659 (2025).

28. Onyeaghala, C. & Iroezindu, M. The 2024-2025 upsurge of mpox in Africa: another opportunity to accelerate global solidarity for a neglected disease. BMJ Glob Health 10, (2025).

29. reliefweb. Cholera and Mpox cases increasing dangerously in DRC as aid cuts push health systems to near-collapse. https://reliefweb.int/report/democratic-republic-congo/cholera-and-mpox-cases-increasing-dangerously-drc-aid-cuts-push-health-systems-near-collapse (2025).

30. Leroy, E. M. et al. Fruit bats as reservoirs of Ebola virus. Nature 438, 575–576 (2005).

31. De Nys, H. M. et al. Survey of Ebola Viruses in Frugivorous and Insectivorous Bats in Guinea, Cameroon, and the Democratic Republic of the Congo, 2015-2017. Emerging Infect. Dis. 24, 2228–2240 (2018).

32. Leendertz, S. A. J., Gogarten, J. F., Düx, A., Calvignac-Spencer, S. & Leendertz, F. H. Assessing the evidence supporting fruit bats as the primary reservoirs for ebola viruses. Ecohealth 13, 18–25 (2016).

33. Koch, L. K., Cunze, S., Kochmann, J. & Klimpel, S. Bats as putative Zaire ebolavirus reservoir hosts and their habitat suitability in Africa. Sci. Rep. 10, 14268 (2020).

34. Pigott, D. M. et al. Mapping the zoonotic niche of Ebola virus disease in Africa. eLife 3, e04395 (2014).

35. Pigott, D. M. et al. Updates to the zoonotic niche map of Ebola virus disease in Africa. eLife 5, (2016).

36. Lee-Cruz, L. et al. Mapping of Ebola virus spillover: Suitability and seasonal variability at the landscape scale. PLoS Negl. Trop. Dis. 15, e0009683 (2021).

37. Vizy, E. K., Manoj, H. & Cook, K. H. Is the Climate of the Congo basin Becoming Less Able to Support a Tropical Forest Ecosystem? J. Clim. 36, 8171–8193 (2023).

38. Didier, L. B., Jude, L. L., Franck, E. I., Wang, H. & Wang, X.-L. Spatial modeling and ecological suitability of Ebola virus disease in Africa. PLoS ONE 20, e0329763 (2025).

39. Kurpiers, L. A., Schulte-Herbrüggen, B., Ejotre, I. & Reeder, D. M. Bushmeat and Emerging Infectious Diseases: Lessons from Africa. in Problematic Wildlife: A Cross-Disciplinary Approach (ed. Angelici, F. M.) 507–551 (Springer International Publishing, 2016). doi:10.1007/978-3-319-22246-2_24.

40. Olivero, J. et al. Recent loss of closed forests is associated with Ebola virus disease outbreaks. Sci. Rep. 7, 14291 (2017).

41. Morton, O. et al. Mining triggers extensive additional deforestation in sub-Saharan Africa. Nature 654, 971–977 (2026).

42. Sovacool, B. K. The precarious political economy of cobalt: Balancing prosperity, poverty, and brutality in artisanal and industrial mining in the Democratic Republic of the Congo. The Extractive Industries and Society 6, 915–939 (2019).

43. Judson, S. D. & Munster, V. J. The multiple origins of ebola disease outbreaks. J. Infect. Dis. 228, S465–S473 (2023).

44. Guéguen, M., Blancheteau, H., Lemaire-Patin, R. & Thuiller, W. biomod2: Ensemble Platform for Species Distribution Modeling. (R, 2026).

45. GBIF.org. GBIF Occurrence Download. 10.15468/dl.r7pbry.

46. GBIF.org. GBIF Occurrence Download. 10.15468/dl.s7at67.

47. GBIF.org. GBIF Occurrence Download. 10.15468/dl.mv2f97.

48. ACR. African Chiroptera Report 2025. (2025).

49. Fick, S. E. & Hijmans, R. J. WorldClim 2: new 1-km spatial resolution climate surfaces for global land areas. Int. J. Climatol. 37, 4302–4315 (2017).

50. Tuanmu, M.-N. & Jetz, W. A global, remote sensing-based characterization of terrestrial habitat heterogeneity for biodiversity and ecosystem modelling. Global Ecology and Biogeography 24, 1329–1339 (2015).

51. Zanaga, D. et al. ESA WorldCover 10 m 2021 v200. 10.5281/zenodo.7254221 (2022).

52. Centre for Disease Prevention and Control. History of Ebola Outbreaks. https://www.cdc.gov/ebola/outbreaks/index.html.

53. Soberon, J. & Peterson, A. T. Interpretation of models of fundamental ecological niches and species’ distributional areas. Biodiv. Inf. 2, (2005).

54. The Malaria Atlas Project. EVI: Malaria Atlas Project Gap-Filled Enhanced Vegetation Index (Annual 1km). Earth Engine Data Catalog https://developers.google.com/earth-engine/datasets/catalog/projects_malariaatlasproject_assets_EVI_v061_1km_Annual.

55. Wan, Z., Hook, S. & Hulley, G. MODIS/Terra Land Surface Temperature/Emissivity Monthly L3 Global 0.05Deg CMG V061 [Dataset]. NASA Land Processes Distributed Active Archive Center 10.5067/MODIS/MOD11C3.061 (2021).

56. Jarvis, A., Reuter, H. I., Nelson, A. & Guevara, E. Hole-filled SRTM for the globe Version 4.

57. McNally, A. et al. A land data assimilation system for sub-Saharan Africa food and water security applications. Sci. Data 4, 170012 (2017).

58. Pesaresi, M. GHS-BUILT-S R2023A - GHS built-up surface grid, derived from Sentinel2 composite and Landsat, multitemporal (1975-2030). European Commission, Joint Research Centre [Dataset] http://data.europa.eu/89h/9f06f36f-4b11-47ec-abb0-4f8b7b1d72ea.

59. Code for Africa (CfA). Rwanda ICGLR Database of Mining Sites 2015. openAFRICA https://open.africa/dataset/rwanda-database-of-mining-sites-2015.

60. Armed Conflict Location & Event Data. Armed Conflict Location & Event Data Project (ACLED). https://acleddata.com/.

61. Jagadesh, S., Zhao, C., Mulchandani, R. & Van Boeckel, T. P. Mapping global bushmeat activities to improve zoonotic spillover surveillance by using geospatial modeling. Emerging Infect. Dis. 29, 742–750 (2023).

62. Hansen, M. C. et al. High-resolution global maps of 21st-century forest cover change. Science 342, 850–853 (2013).

63. Hirzel, A. H., Le Lay, G., Helfer, V., Randin, C. & Guisan, A. Evaluating the ability of habitat suitability models to predict species presences. Ecol. Modell. 199, 142–152 (2006).

64. Di Cola, V. et al. ecospat: an R package to support spatial analyses and modeling of species niches and distributions. Ecography 40, 774–787 (2017).

